# Causal Effects Contributing to Elevated Metabolic Power During Walking in Children Diagnosed with Cerebral Palsy

**DOI:** 10.1101/2022.01.26.22269878

**Authors:** Pavreet K. Gill, Katherine M. Steele, J. Maxwell Donelan, Michael H. Schwartz

## Abstract

Metabolic power (net energy consumed while walking per unit time) is, on average, two-to-three times greater in children with cerebral palsy (CP) than their typically developing peers, contributing to greater physical fatigue, lower levels of physical activity and greater risk of cardiovascular disease. The goal of this project was to identify the total causal effects of clinical factors that may contribute to high metabolic power demand in children with CP.

We included children who 1) visited Gillette Children’s Specialty Healthcare for a quantitative gait assessment after the year 2000, 2) were formally diagnosed with CP, 3) were classified as level I-III under the Gross Motor Function Classification System and 4) were 18 years old or younger. We created a structural causal model that specified the assumed relationships of a child’s gait pattern (i.e., gait deviation index, GDI) and common impairments (i.e., dynamic and selective motor control, strength, and spasticity) with metabolic power. We estimated causal effects using Bayesian additive regression trees, adjusting for factors identified by the causal model.

There were 2157 children who met our criteria. We found that a child’s gait pattern, as summarized by the GDI, affected metabolic power approximately twice as much as the next largest contributor. Selective motor control, dynamic motor control, and spasticity had the next largest effects. Among the factors we considered, strength had the smallest effect on metabolic power.

Our results suggest that children with CP may benefit more from treatments that improve their gait pattern and motor control than treatments that improve spasticity or strength.

## 1 Introduction

The net metabolic power required during walking is, on average, two-to-three times greater in children with cerebral palsy (CP) than their typically developing peers (Campbell and Ball, 1978; Rose et al., 1990). As a result, walking is as difficult as performing moderate- to vigorous-intensity exercise (Balemans et al., 2017). Net metabolic power during walking (metabolic power) is the metabolic energy consumed while walking minus the metabolic energy consumed while resting per unit time. High metabolic power contributes to greater physical fatigue (Jahnsen et al., 2007; Balemans et al., 2017), lower physical activity (Carlon et al., 2013; Ryan et al., 2014), increased risk of cardiovascular disease (Hammam et al., 2021), and reduced participation in school and the community (Verschuren et al., 2012). Reducing or preventing fatigue is one of the top research priorities of the CP community (Gross et al., 2018) and reducing metabolic power is a mechanism by which to do so.

Due to brain injury near the time of birth, children with CP exhibit various neurological impairments that may contribute to high metabolic power. Spasticity is present in up to 91% of children with CP (Odding et al., 2006) and clinicians often believe it causes high metabolic power via inappropriate or prolonged muscle contraction and co-contraction of agonists and antagonists. However, several recent studies have indicated that spasticity reduction does not cause meaningful reductions in metabolic power during walking (Ubhi et al., 2000; Thomas et al., 2004; Bjornson et al., 2007; Munger et al., 2017; Zaino et al., 2020). Reduced metabolic power following spasticity treatment in observational settings likely arises from other factors such as aging. In addition to spasticity, muscle weakness and poor motor control are also common neurological impairments in CP. Both may contribute to high metabolic power via inefficient muscle activation and altered gait mechanics (Damiano et al., 2000; Rose and McGill, 2007; Bennett et al., 2012; Steele et al., 2015; Schwartz et al., 2016), but they are inextricably linked, so it is challenging to isolate their effects. Conner et al. (2020) recently showed that resistance training via ankle exoskeletons significantly improved muscle strength and reduced metabolic power in children with CP. Further, their exoskeleton training also caused significant improvements in motor control and walking mechanics (Conner et al., 2021). Prior studies of resistance training have demonstrated mixed results (MacPhail and Kramer, 1995; Damiano and Abel, 1998; Eagleton et al., 2004; Ryan et al., 2020), suggesting that muscle weakness may also be a cause of high metabolic power. Understanding the relative effects of strength, motor control, and other factors on high metabolic power in CP is challenging, but important to understand and reduce these demands.

In addition to neurological impairments, children with CP also exhibit atypical gait kinematics that may contribute to high metabolic power. Bony malalignments are common in CP and contribute to altered gait kinematics. Many children undergo orthopedic surgery to correct these malalignments and display significantly improved gait kinematics post-treatment. However, no controlled study has directly investigated whether changes in gait kinematics after surgery – in contrast to other factors such as aging – cause significant reductions in metabolic power. For example, McMulkin et al., (2016) assessed surgical outcomes for children undergoing multi-level surgery with and without a femoral derotation osteotomy. In this study, treatment and control groups were matched except for the inclusion of a femoral derotation osteotomy. McMulkin’s study showed that a femoral derotation osteotomy results in slightly better outcomes after surgery (McMulkin et al., 2016), but was not able to show that better gait kinematics cause lower metabolic power since both groups underwent near-identical treatments and experienced similar changes in gait kinematics. Similar studies provide further insights into how gait kinematics and metabolic power change after surgery (Thomas et al., 2004; Wren et al., 2013), but cannot address this central question of whether improved kinematics lowers the metabolic power required to walk.

Currently, experimental data does not clearly refute or confirm various factors as causes of high metabolic power in CP. The gold standard for making causal inferences is to perform controlled experiments, where all other factors except the factor of interest are kept constant. However, controlled experiments are not always possible. For example, it is not possible to keep kinematics and kinetics constant while changing muscle spasticity. In place of experimental data, we may be able to analyze the natural variability present in observational data to understand what factors are responsible for high metabolic power and their relative effect sizes.

Researchers often hesitate to make causal inferences from observational data due to confounding. Two things are needed to infer causation – the “cause” must occur before the “effect” (temporal priority) and all other “potential causes” (confounders) must be controlled for. Confounding is common in observational studies since data are collected in uncontrolled environments. We can reduce confounding by statistically controlling (adjusting) for factors, but we may induce bias if we fail to do so correctly. MacWilliams and colleagues have shown that without appropriate adjustment, linear regression analysis can greatly over or underestimate effect sizes for factors causing function limitations in CP (MacWilliams et al., 2020). If we can correctly identify confounders, causal inferences are possible (Pearl, 1995).

Causal models provide a systematic approach to identifying confounders. One type of causal model is the structural causal model, often depicted as a directed acyclic graph. A directed acyclic graph is a graphical representation of causal relationships that can be queried to determine possible confounders (Pearl, 1995). Directed acyclic graphs are easier to use and less likely to fail at identifying confounders compared to traditional methods (Hernán and Robins, 2020), enabling causal inferences from observational data.

In the study described here, we (1) propose a causal model for metabolic power in CP as a function of gait kinematics, selective motor control, dynamic motor control, strength, and spasticity, and (2) compute the total causal effect of each factor on metabolic power with large scale data. We decided to investigate these five factors as they are commonly measured or treated and likely contribute to high metabolic power in children with CP. We used directed acyclic graphs to form our causal model and identify confounders. Then, we used Bayesian additive regression trees (BART) to compute the causal effects of these five factors, including model-identified confounders in the analysis to adjust for their effects.

## 2 Materials and Methods

### 2.1 Participants

We obtained approval for this research from the Research Ethics Board (REB) at Simon Fraser University.

We retrospectively analyzed data collected from 6220 children seen between the years 2000 and 2020 at the Center for Gait and Motion Analysis at Gillette Children’s Specialty Healthcare. We selected children who met the following criteria: formally diagnosed with CP; classified as level I, II, or III under the Gross Motor Function Classification System (GMFCS); 18 years or younger; and had undergone a quantitative 3D gait assessment and a 6-minute walking metabolic assessment. Many children had visited the lab more than once. To avoid pseudoreplication, we selected the visit with the least missing data.

### 2.2 Data Collection

Spasticity was measured by a trained physical therapist using the Ashworth Scale (Bohannon and Smith, 1987; Pandyan et al., 1999). The Ashworth scale is a 5-point scale defined by the following: (1) no increase in tone, (2) slight increase in tone, (3) more marked increase in tone, (4) considerable increase in tone, and (5) rigidity. Six muscle groups were assessed bilaterally: hip flexors, hip adductors, rectus femoris, hamstrings, plantarflexors, and tibialis posterior. We calculated a summary spasticity score by applying polychoric principal component analysis to the bilateral measurements from these six muscles for individuals with complete data (Rozumalski and Schwartz, 2009; Zaino et al., 2020). We used polychoric principal component analysis since standard principal component analysis produces biased estimates with categorical data (Kolenikov and Angeles, 2009).

Strength was measured by a physical therapist for the hip flexors, hip adductors, rectus femoris, hamstrings, plantarflexors, and tibialis posterior. This was measured on a 5-point scale where 1 is defined as a ‘visible or palpable contraction (no range of motion)’ and 5 is defined as ‘full range of motion against gravity’. Again, we calculated a summary strength score using polychoric principal component analysis, which included measurements from both lower limbs for individuals with complete data (12 measurements total).

Selective motor control (SMC) was measured by a physical therapist using a 3-point scale defined by the following: (0) very little or no control of a single joint voluntary movement, (1) impaired voluntary movement at a single joint, and (2) good voluntary movement at a joint. The physical therapist measured SMC for hip flexion, hip adduction, knee extension, knee flexion, plantarflexion, and posterior tibialis. Again, we calculated a summary SMC score with the same methods used to calculate summary spasticity and strength scores.

Patients underwent a quantitative 3D gait analysis with electromyography of the anterior tibialis, lateral and medial hamstrings, gastroc-soleus, and rectus femoris muscles. Using the motion capture and electromyographic data, respectively, we calculated gait deviation index (GDI) (Schwartz and Rozumalski, 2008)and dynamic motor control (DMC) (Shuman et al., 2019) scores for each child.

Breath-to-breath oxygen consumption was measured during six minutes of overground, barefoot walking around a rectangular 80-meter track (Ultima CPX, Medical Graphics Corporation, St. Paul, MN, USA). Breath-to-breath oxygen consumption was also measured during the 10 minutes of reclining rest that preceded the walking session. The average steady state value was calculated for both periods according to the method proposed by (Schwartz, 2007). Oxygen consumption per unit time was converted to power using the conversion rate of 20.1 Joules/mL O2 (Lusk, 1924; Taylor and Heglund, 1982). We then calculated net metabolic power by subtracting resting from walking values. We also calculated average walking speed during the trial and recorded patient age, height, mass, and sex.

### 2.3 Causal Model

Structural causal models are visual representations of causal assumptions that we can systematically test for plausibility and query to identify confounders. Nodes represent variables, and arrows drawn (or excluded) between variables indicate causal relationships (or lack thereof) between them. The absence or presence of arrows can be used to determine conditional independencies implied by the causal model. By checking these independencies, we can test the plausibility of a model. And, just as we can test a model’s plausibility, we can also determine from the model what variables are potential confounders (see Greenland & Pearl (2017) for a detailed explanation). We can then statistically adjust for these potential confounders in our computational analysis to reduce potential bias (Pearl, 1995). It may be necessary to adjust for more than one factor. An adjustment set is the minimum set of factors to adjust for when computing the causal effect of X on Y (Pearl, 1995).

We define causal effects as the *total* effect of X on Y. This includes both the direct (unmediated) and indirect (mediated) effects of X on Y. We focused on the total effect of each factor as treatments in CP rarely affect a single factor due to their inter-relatedness. For example, strength may directly affect metabolic power, but it may also indirectly affect metabolic power via changes in kinematics.

We built a causal model for this study that represents our hypotheses and assumptions about the mechanisms contributing to metabolic demand during walking (Figure 1). We created and tested the proposed model using the *dagitty* package in R (Textor et al., 2016). From our causal model, we determined adjustment sets that we used in our data analysis to compute the causal effects of each factor on metabolic power.

**Figure 1.**
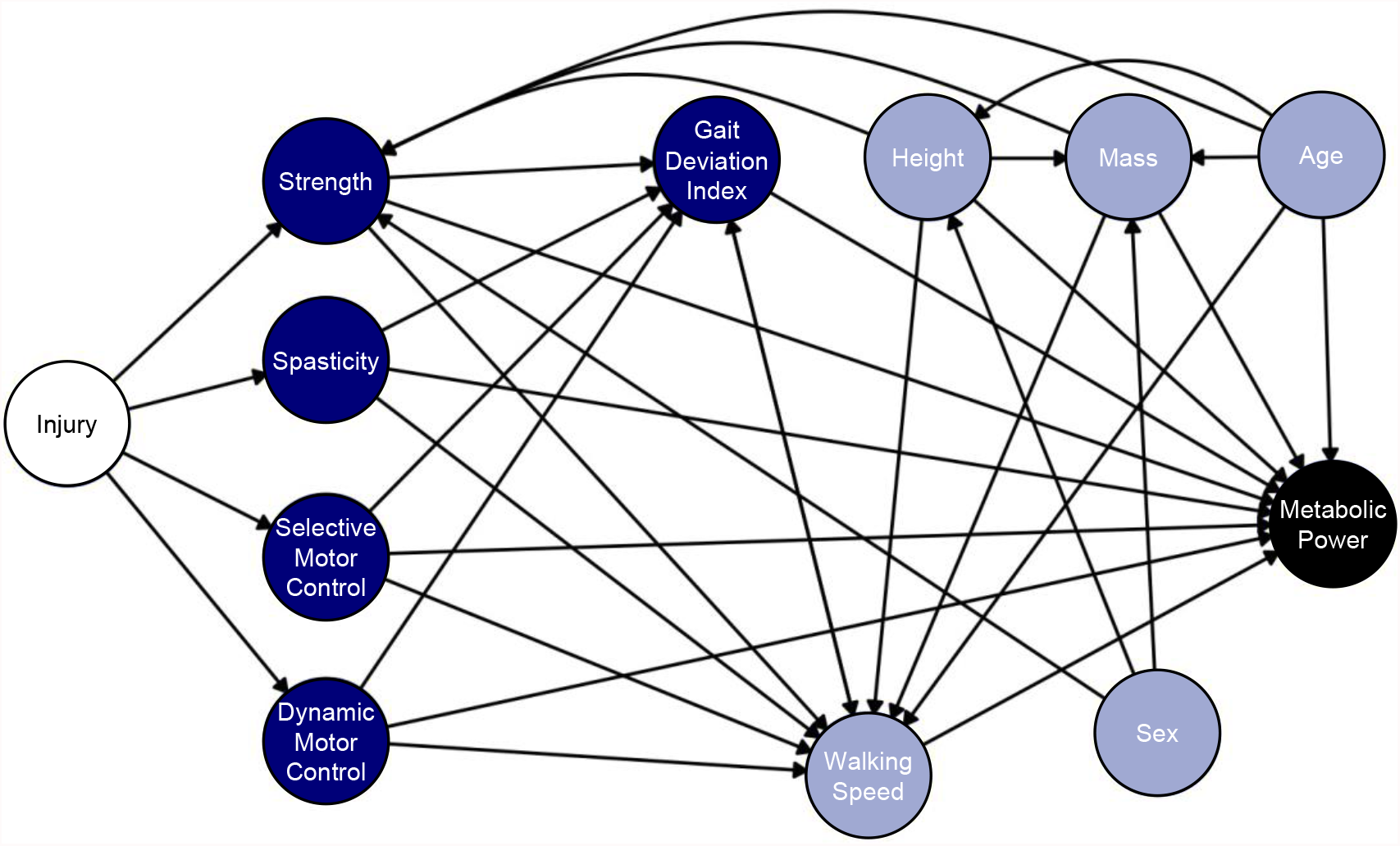
Causal model (portrayed as a directed acyclic graph) of the potential contributors to high metabolic power during walking in children with cerebral palsy (CP). The initial brain injury is colored white to represent an unmeasured factor. Despite being unmeasured, including the initial brain injury helps to simplify the complex relationship between neurological impairments in CP. Blue nodes are factors often treated and measured in the clinic (neurological and physical impairments), so understanding their effects offers a better understanding of their importance in treating metabolic power. Metabolic power, colored in black, is the outcome of interest. Gray variables are potential confounders.

### 2.4 Data Analysis

After obtaining adjustment sets from our causal model, we used the Bayesian additive regression trees (BART) method (Chipman et al., 2010) to compute the causal effects of GDI, DMC, SMC, strength, and spasticity on metabolic power. Since BART is a regression-based approach, including factors from each adjustment set as predictors in the BART model adjusted for their effects and reduced confounding. A causal model identifies sources of bias, but it does not provide information regarding the strength or direction of a causal relationship. Thus, it was necessary to use regression analysis (i.e., BART) to compute effect sizes.

The BART method is an excellent option to identify the magnitude of causal effects from observational data in CP. The BART method demonstrates superior predictive abilities against other common methods (Dorie et al., 2019) and has been successfully used in the causal inference of observational data (Hill, 2011). Many observational studies make linear assumptions. CP is a complex condition, and although linear or multiple linear regression are convenient and easy-to-understand tools, it is unwise to assume linearity between factors. For example, as a child’s tibial torsion increases (either internally or externally), we can expect their walking pattern to deteriorate, resulting in a U-shaped relationship. Besides its predictive prowess, BART is a non-parametric tool that a) uses Bayesian methods to produce estimates with honest uncertainty bounds in its predictions, b) can handle large numbers of predictor variables of various types (scale, ordinal, categorical), c) requires little-to-no hand-tuning, and d) does not require investigator involvement to determine the shape of the response surface (Hill, 2011). In addition, BART natively handles missing data (increasing sample representativeness) and outperforms imputation methods when dealing with data not missing at random (Kapelner and Bleich, 2015).

To visualize and interpret the causal effects specified by our BART models, we utilized accumulated local effects plots (Apley and Zhu, 2020). These plots show the average change in the response variable (i.e., metabolic power) due to changes in the predictor variable (i.e., GDI, DMC, SMC, strength, or spasticity). Machine learning algorithms, such as BART, are harder to interpret than linear regression because they use more complex (and often hidden) functions. By visualizing the results, we could interpret the causal effects specified by our model. Accumulated local effects plots, in particular, are unbiased when features are highly correlated compared to other visualization methods, such as partial dependence plots (Apley and Zhu, 2020).

We used the R package *bartMachine* (Kapelner and Bleich, 2015) to create our BART models. Following hyperparameter optimization, the model settings were: *num_trees = 50, k = 5, nu = 3, q = 0*.*99, use_missing_data = TRUE*. For replicability, we set the seed of each model as 42. To visualize the causal effects of each factor in the form of accumulated local effects plots, we used the R package *ALEPlot* (Apley, 2018). From each plot, we approximated effect sizes using the range of the middle 95th percentile of the sample.

## 3 Results

### 3.1 Participants

We analyzed data from 2157 children who met our inclusion criteria (Table 1). Of those children, 32.1% were missing DMC scores, 18.3% were missing spasticity scores, 17.7% were missing strength scores, 17.5% were missing SMC scores, and 0.6% were missing GDI scores.

**Table 1.** Participant Characteristics.

|  | Total | GMFCS I | GMFCS II | GMFCS III |
| --- | --- | --- | --- | --- |
| n (%) | 2157 | 662 (30.7%) | 1000 (46.4%) | 495 (22.9%) |
| Sex = male (%) | 1212 (56.2%) | 375 (56.6%) | 552 (55.2%) | 285 (57.6%) |
| Age (median (SD)) | 9.3 (3.7) | 10.1 (3.5) | 8.8 (3.7) | 8.2 (3.7) |
| Height (median (SD)) | 129.5 (20.3) | 136.5 (18.8) | 127.5 (20.4) | 119.9 (19.4) |
| Mass (median (SD)) | 26.9 (15.8) | 31.4 (16.1) | 26.5 (15.9) | 23.5 (14.6) |
| GDI (median (SD)) | 71.1 (12.5) | 80.0 (10.3) | 70.9 (11.1) | 60.7 (9.2) |
| DMC (median (SD)) | 83.3 (9.9) | 90.3 (8.0) | 81.7 (8.7) | 74.1 (8.3) |
| SMC (median (SD)) | 0.25 (1.1) | 0.88 (0.7) | 0.13 (1.0) | -1.11 (1.0) |
| Spasticity (median (SD)) | -0.25 (1.1) | -0.52 (0.7) | -0.16 (1.1) | 0.36 (1.3) |
| Strength (median (SD)) | 0.16 (1.1) | 0.85 (0.7) | 0.08 (0.9) | -1.15 (0.9) |
| Met. Power (median (SD)) | 124.2 (75.3) | 119.9 (66.9) | 127.5 (77.2) | 120.1 (81.2) |

Height is described in cm. Mass is described in kg. SMC, spasticity, and strength are scores derived from polychoric principal component analysis.

### 3.2 Outcomes of Causal Model

Our model satisfied all implied conditional independencies. The conventional cut-off for independence is ±0.3. All partial correlation coefficients were smaller than ±0.2, suggesting that the observed data was consistent with the proposed relationships in our causal model (i.e., our model was plausible). From our model, we obtained sufficient adjustment sets for all variables (Table 2). Adjustment sets were the same for DMC, SMC, spasticity and strength.

**Table 2.** Adjustment sets necessary to minimize confounding bias.

|  | Adjustment Set |
| --- | --- |
| GDI | Walking speed, DMC, SMC, spasticity, strength, age, height, mass |
| DMC | SMC, spasticity, strength, age, height mass |
| SMC | DMC, spasticity, strength, age, height, mass |
| Spasticity | DMC, SMC, strength, age, height, mass |
| Strength | DMC, SMC, spasticity, age, height, mass |

### 3.3 Outcomes of BART

The purpose of this project was to determine the causal effects of GDI, DMC, SMC, strength and spasticity on metabolic power. Based on the adjustment sets indicated by our assumed causal model, we only required two BART models: one to compute the effects of GDI (r^2^ = 0.81, rmse = 32.96) and one to compute the effects of DMC, SMC, strength and spasticity (r^2^ = 0.72, rmse = 39.48). From the accumulated local effects plots (Figure 2), we approximated effect sizes using the range of the middle 95th percentile of the sample (Figure 3). GDI had approximately a two-fold effect on metabolic power than DMC, SMC and spasticity. Strength had the smallest effect on metabolic power. These results indicate that gait kinematics had the largest effect on high metabolic power.

**Figure 2.**
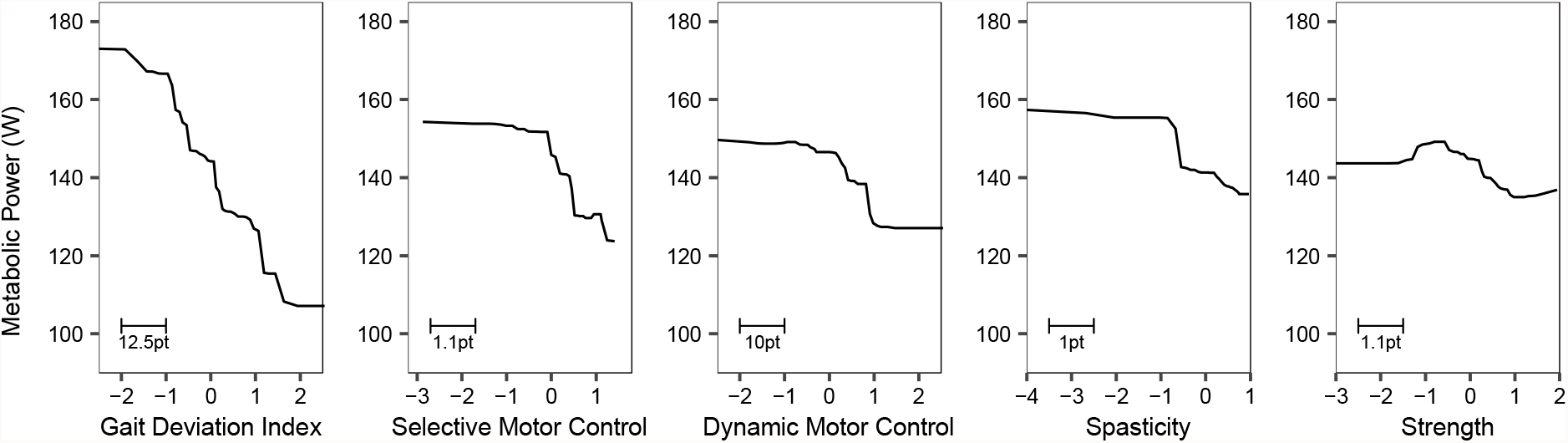
Accumulated local effects plot of the total causal effects of gait deviation index, selective motor control, dynamic motor control, spasticity, and strength on metabolic power with 95% bootstrapped confidence intervals. These plots represent the average change in metabolic power that can be expected with a change in the x-variable. X-axis variables are normalized as z-scores, where increasing or more positive scores indicate a lesser severity of impairment (i.e., more typical gait, more coordinated, less spastic, stronger). Rug plots along the x-axis display the distribution of scores for each impairment. The bottom right scale indicates how large a single standard deviation is with respect to original units.

**Figure 3.**
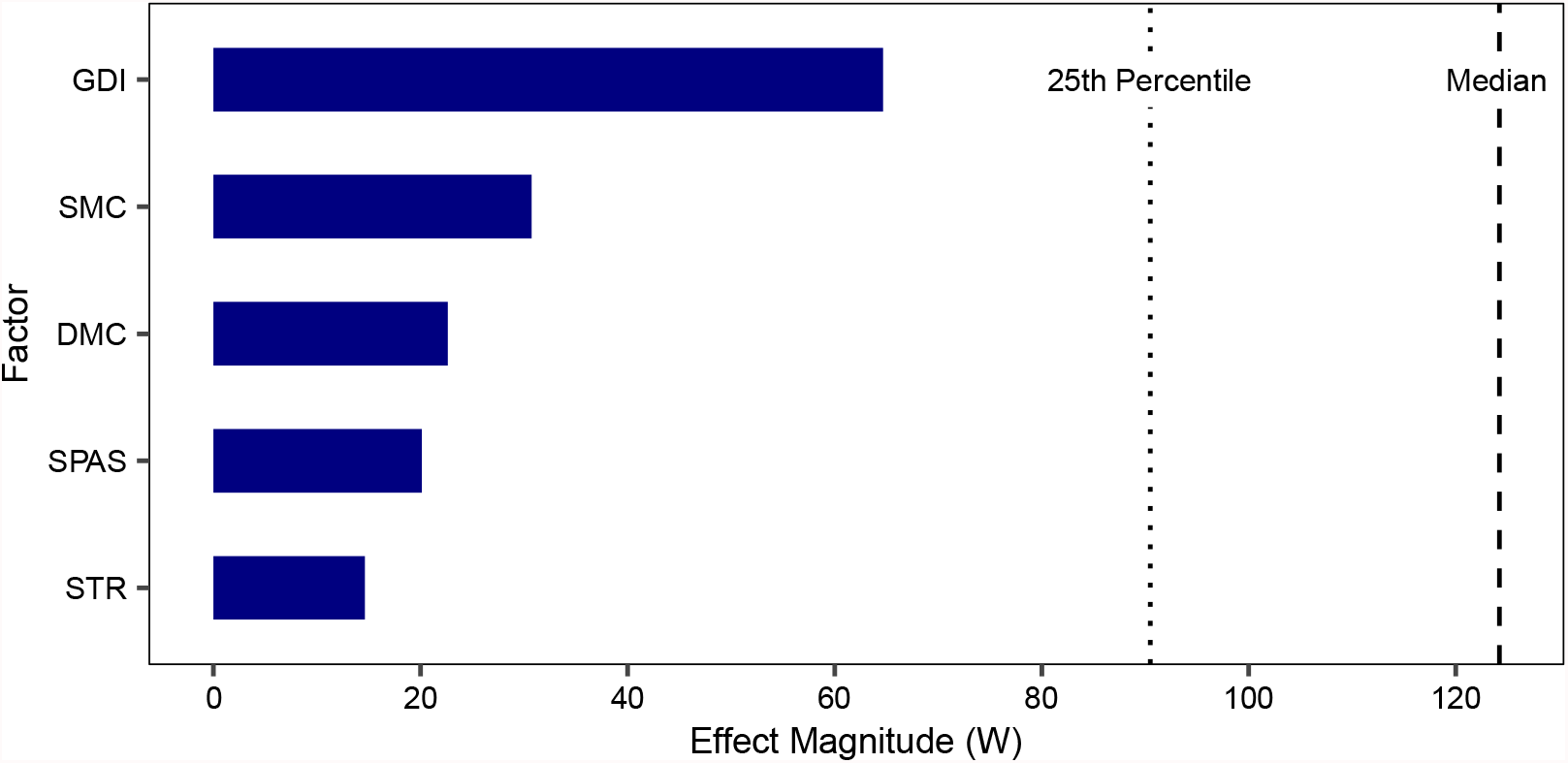
The range of the causal effect for each factor for the middle 95th percentile. The dotted and dashed lines show the 25th percentile and median metabolic power of children included in this study. The total effect of GDI is approximately twice that of SMC and DMC, and more than thrice that of spasticity or strength.

The GDI had the steepest curve (Figure 2), on average, suggesting that changing GDI can elicit the largest changes in metabolic power. Most of the factors shared a near-linear or sigmoidal relationship with metabolic power. However, strength and metabolic power shared an almost inverted-U relationship. Regardless of shape, most factors plateaued at the extremes of each plot. These plateaus suggest that after a certain level of impairment, metabolic power may not increase or decrease considerably. So, children with mild or very severe level impairments may experience limited reductions in metabolic power following treatment. For example, children with largely in-tact dynamic motor control may experience limited changes in metabolic power due to ceiling effects whereas, children with severely compromised DMC may experience limited changes in metabolic power due to movement limitations.

## 4 Discussion

Reducing metabolic power is a means of reducing fatigue and facilitating physical activity in children with CP. Our analysis indicates that gait kinematics (as quantified by GDI) had the largest causal effect on metabolic power, more than twice as large as the other factors. The effects of SMC, DMC and spasticity had the next largest contributions to metabolic power. Strength had the smallest effect. These results suggest that improving gait kinematics may elicit reductions in metabolic power that are more likely to be clinically meaningful.

On average, children with CP need to reduce their metabolic power by 50% to achieve values within the typical range. Such a reduction may not be possible, but a 10% reduction in metabolic power is still considered clinically meaningful (Oeffinger et al., 2008). This value can be used to interpret the observed effects in this study.

We cannot perform randomized control trials, but we must assess the order of magnitude of our results to either confirm or refute our findings. The model-predicted changes in metabolic power following changes in GDI are comparable to experimental studies. In the study by McMulkin et al., children (GMFCS level I/II) who underwent surgical intervention with a femoral derotation osteotomy improved their GDI by ∼13 points and their net oxygen cost (net volume of oxygen consumed per unit distance) by ∼15% (McMulkin et al., 2016). According to our results, metabolic power would decrease by 12-27W if GDI increased by 13 points. For a child with median metabolic power in our sample (124W), this would be equivalent to a 10-22% decrease in metabolic power. The values seen by McMulkin et al. fall within this range. In another study, children with CP improved their GDI by approximately five points and their net oxygen cost by 2.5% one year following surgical intervention (Wren et al., 2013). According to our results, metabolic power would change by 2-13% for a child with the median metabolic power if the GDI increases by five points. While our estimates seem high, Wren et al. may have found insignificant results because their sample included children with more severe CP (i.e., GMFCS level IV). Children with more severe impairments (i.e., GMFCS level III and IV) have significantly greater metabolic power during walking (Bolster et al., 2017). So, even if they experience similar *absolute* reductions in metabolic power, *relative* reductions in metabolic power may not be as significant because metabolic power is higher overall. This may be why Wren et al. saw lower changes than those predicted by our results. Further, McMulkin et al. only saw significant reductions in metabolic power for children with GMFCS level I/II despite greater *absolute* reductions in metabolic power for children with GMFCS level III. Thus, treatments that produce greater changes in gait kinematics may yield more significant results for children with lower metabolic power when we consider relative rather than absolute changes.

This research highlights how myriad factors impact metabolic power in CP, which can provide numerous pathways for potentially improving walking power. Our model suggests that changes in GDI and motor control affect metabolic power. Interventions that can improve these factors may improve metabolic power. This was recently observed in a pilot study by Conner et al. A small group of children with CP improved DMC by 7% and reduced metabolic power by 29% after resistance training with an ankle exoskeleton (Conner et al., 2021). According to our results, metabolic power should decrease by 1-11% for a 7% improvement in DMC. These estimates are lower than Conner et al., but Conner et al. have also shown that their exoskeleton training results in mechanically more efficient gait (Conner et al., 2021) and significantly greater muscle strength (Conner et al., 2020). Since our model does not include a metric of gait kinetics, this could have affected our causal effect estimates. In addition, a large proportion of children in our sample (31.2%) are missing DMC scores. Although BART natively handles missing data, greater missing data increases model uncertainty. Both unexplained factors and model uncertainty might explain why our estimates are lower. We estimate SMC has similar effects on metabolic power as DMC. SMC and DMC are only moderately correlated (r = 0.54), which suggests that although both are related, they also explain different aspects of motor control in CP. Thus, the treatment of children with CP should include both selective and dynamic motor control training.

Despite modest total effects, spasticity itself may not be a large contributor to high metabolic power. In a study by Zaino et al. (2020), children who underwent selective dorsal rhizotomy experienced a 0.9 point reduction in lower body spasticity (approximately a change of one level out of four typically seen) and a 12% reduction in metabolic power, on average. For the same reduction in spasticity, our results indicate that metabolic power should decrease by 2-14% for a child with the median metabolic power. Our estimates capture the values found by Zaino et al. Similar to Zaino’s study, our results reflect the total effect of spasticity on metabolic power. Total effects include indirect effects, such as those associated with changes to gait kinematics, so spasticity alone may not be an important determinant of metabolic power. To further support this point, Zaino et al. found changes in metabolic power were not significantly different when compared to a control group, and when we calculate the direct effect of spasticity, it is only a third of its total effect. Therefore, other factors may be responsible for meaningful reductions in metabolic power following spasticity reduction.

Compared to the other variables assessed in this study, strength has smaller causal effects and training strength is not likely to cause meaningful reductions in metabolic power. With a single standard deviation change in strength, our model predicts that metabolic power changes by 1-7% for the average child in our sample. While this may seem like a small reduction, it is not entirely unexpected. As a result of strength training, children with CP often walk faster and for longer distances (Damiano and Abel, 1998; Eagleton et al., 2004). Since walking faster ultimately increases metabolic power, it would explain why strength has a smaller total effect on metabolic power. Interestingly, strength shares a slight inverted-U-shaped relationship with metabolic power. Weaker individuals may experience increased metabolic power after strength training because they can walk longer and faster. And stronger individuals may experience decreased metabolic power after strength training because their motor control has improved along with their strength, so they can walk more efficiently. Other studies have also speculated that different responses to strength training arise due to changes in walking speed and mechanical efficiency (Damiano and Abel, 1998; Eagleton et al., 2004). But, although metabolic power may not change drastically, strength training is still important for improving other functional outcomes important to children with CP (Conner et al., 2020).

We did not normalize or non-dimensionalize metabolic power. We made this choice to simplify our causal model and avoid spurious relationships caused by imperfect normalization. Most non-dimensionalization schemes assume a linear dependence of metabolic power on mass. While this assumption holds true in typically developed populations, it may not apply to children with CP. A study by Plasschaert et al. simulated weight gain during walking in children with and without CP – they noticed that mass normalization was not as effective in removing mass-dependence for the children with CP (Plasschaert et al., 2008). We noticed a similar phenomenon in our data. This suggests that mass and metabolic power share a non-linear relationship in children with CP. If this is true, decreases in mass may elicit greater reductions in metabolic power for a child with CP than a typically developed child. The opposite would also be true, and both would have important implications for weight management in children with CP.

Our model, like all models, requires assumptions. An advantage of structural causal models is that the assumptions are made clear and explicit, and the plausibility of those assumptions is tested. So, while our model may not perfectly represent the causes of high metabolic power in CP, by explicitly specifying our causal assumptions, researchers will be able to compare their results and models. We believe our causal model is a valuable starting point and will lead to experimental research that accepts or rejects hypotheses informed by these results.

Reducing metabolic power can play an important role in reducing or delaying fatigue onset in children with CP and allowing them to be more physically active. Using large-scale data spanning a diverse study sample of widely varying age, GMFCS level, and CP subtype, we sought to understand the causal effects of commonly treated variables (i.e., GDI, SMC, DMC, spasticity, and strength) on metabolic power. Our findings suggest that impaired gait kinematics contribute the most to high metabolic power, followed by SMC and DMC. If the primary goal of treatment is to reduce fatigue, improving motor control and gait mechanics may be the most effective treatment options. Future research should look to perform controlled experiments to test the legitimacy of these findings.

## Data Availability

All data produced in the present study are available upon reasonable request to the authors

## 6 Author Contributions

P.K.G., K.M.S., J.M.D., and M.H.S. conceived and designed the research; P.K.G. analyzed the data, prepared figures, and drafted the manuscript; P.K.G., K.M.S., J.M.D., and M.H.S. interpreted the results, revised the manuscript, and approved the final version of the manuscript.

